# “Like brushing teeth” – Implementation experiences with opt-in, at-home screening for SARS-CoV-2 among schoolchildren in Germany

**DOI:** 10.1101/2021.09.13.21263486

**Authors:** Jonas Wachinger, Maximilian Schirmer, Nicole Täuber, Shannon A. McMahon, Claudia M. Denkinger

## Abstract

**Background:** Over the course of the pandemic, many countries have repeatedly closed schools and shifted students to remote learning. However, evidence for negative mental and physiological health consequences of such measures for students is increasing, highlighting the need for evidence-based recommendations on how to safely reopen schools. This study presents experiences when implementing opt-in, at-home SARS-CoV-2 screening using rapid diagnostic tests (RDTs) to facilitate safe face-to-face-teaching during a pandemic.

**Methods:** We present data form a prospective study implementing an RDT-based screening program at a primary school in southwest Germany. We conducted qualitative in-depth interviews with participating children, parents, and school stakeholders to elicit implementation experiences and screening perception.

**Results:** The screening intervention was highly accepted and appreciated among participants; no positive RDT was reported over the duration of the study. Self-testing at home before coming to school was feasible, but more positive consequences of screening participation (e.g., easing of mask mandates) besides a personal feeling of safety would be appreciated. Participants preferred home-based RDTs over some other measures, particularly mask mandates. Despite the RDTs being licensed as self-tests in Germany, additional training can help avoid mistakes, and ensuring intervention ownership and improving pre-implementation communication can facilitate buy-in.

**Conclusions:** Ag-RDT-based SARS-CoV-2 screening programs relying on self-testing at home proved feasible and accepted among primary school students, parents, and school staff who participated in this study.

**Trial Registration:** DRKS00024845

What is known about the subject

- Efforts to reduce COVID-19-associated school disruption are currently being debated globally as a means to reduce the impact of extended school closures on children’s mental and physiological wellbeing.
- Rapid diagnostic tests (RDTs) for SARS-CoV-2 are reliable and can be performed as self-tests at home.
- Although countries have already introduced RDT-based screening programs to facilitate safe face-to-face teaching, little is known about screening acceptance and experiences.

What this study adds

- Students, parents, and school staff perceive home-based RDT screening as feasible and less disrupting than other protective measures (e.g., mask mandates)
- Implementers should communicate early and clearly, and provide a support system for training, troubleshooting, and in case of positive results
- Concerns remain regarding the fidelity of home-based test performance in cases where students or parents are hesitant, even when testing is compulsory

## INTRODUCTION

To curb infection rates in the context of the COVID-19 pandemic many countries closed primary and secondary schools, and children were shifted to remote learning to minimize risks of viral transmission.[1, 2] However, in light of growing evidence regarding the impact of prolonged school closure for children’s education and mental health and children’s limited impact on viral transmission dynamics, schools were reopened accompanied by the implementation of hygiene measures.[3] An addendum to the toolbox of measures entailed either antigen-based rapid diagnostic tests (RDTs) or pooled Polymerase chain reaction (PCR) testing for SARS-CoV-2.

However, the idea of large-scale screening efforts at schools was criticized both from an epidemiological perspective (regarding imperfect test performance, especially of RDTs), but also as being an unnecessary burden for school children.[4–6] While a study from Great Britain suggests that SARS-CoV-2 protective measures in schools are highly accepted among students and parents,[7] to the best of our knowledge there is no evidence available on the perceptions of and experiences with the implementation of testing for entrance screening in school settings and the effects on compliance with other safety measures (e.g., masks).

An in-depth investigation of school-based testing implementation would facilitate evidence-based recommendations for best practices of entrance screening in schools, not only in the context of this pandemic but also for future public health crises. This study fills a gap in the literature by providing implementation insights regarding RDTs for home-based screening of primary school children in Germany.

## METHODS

We conducted a prospective implementation study to assess experiences with and perceptions of introducing in-home RDT-based screening at a primary school in a peri-urban area of southwestern Germany. Throughout the early pandemic, schools in the region were routinely fully or partly closed, and an increasing number of academics, policymakers, school representatives, and parents argued for schools to be reopened with comprehensive screening approaches complementing other hygiene measures.[8, 9]

Responding to calls for pilot projects testing the feasibility and acceptability of such screening efforts, our study-based screening was initiated in March 2021. Statewide compulsory screening was introduced for schools in April 2021. Figure 1 presents a timeline of study-related processes and the general context.

**Figure 1.**
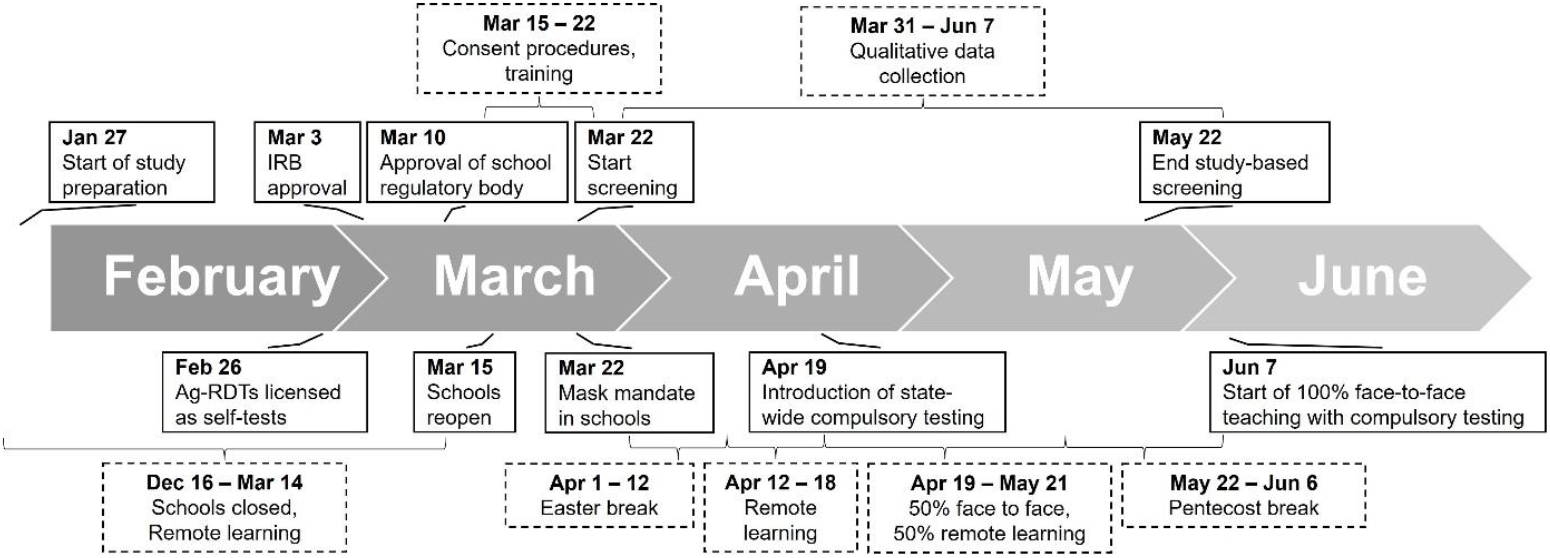
Study processes and the general context.

### Intervention design

The design of the screening intervention was developed in partnership with school stakeholders (Figure 2). For each week of screening, school students and staff members who voluntarily decided to participate in the study received three STANDARD Q COVID-19 Ag Tests (SD Biosensor, Inc. Gyeonggi-do, Korea), an independently validated and WHO-approved SARS-CoV-2 RDT.[10, 11]

**Figure 2.**
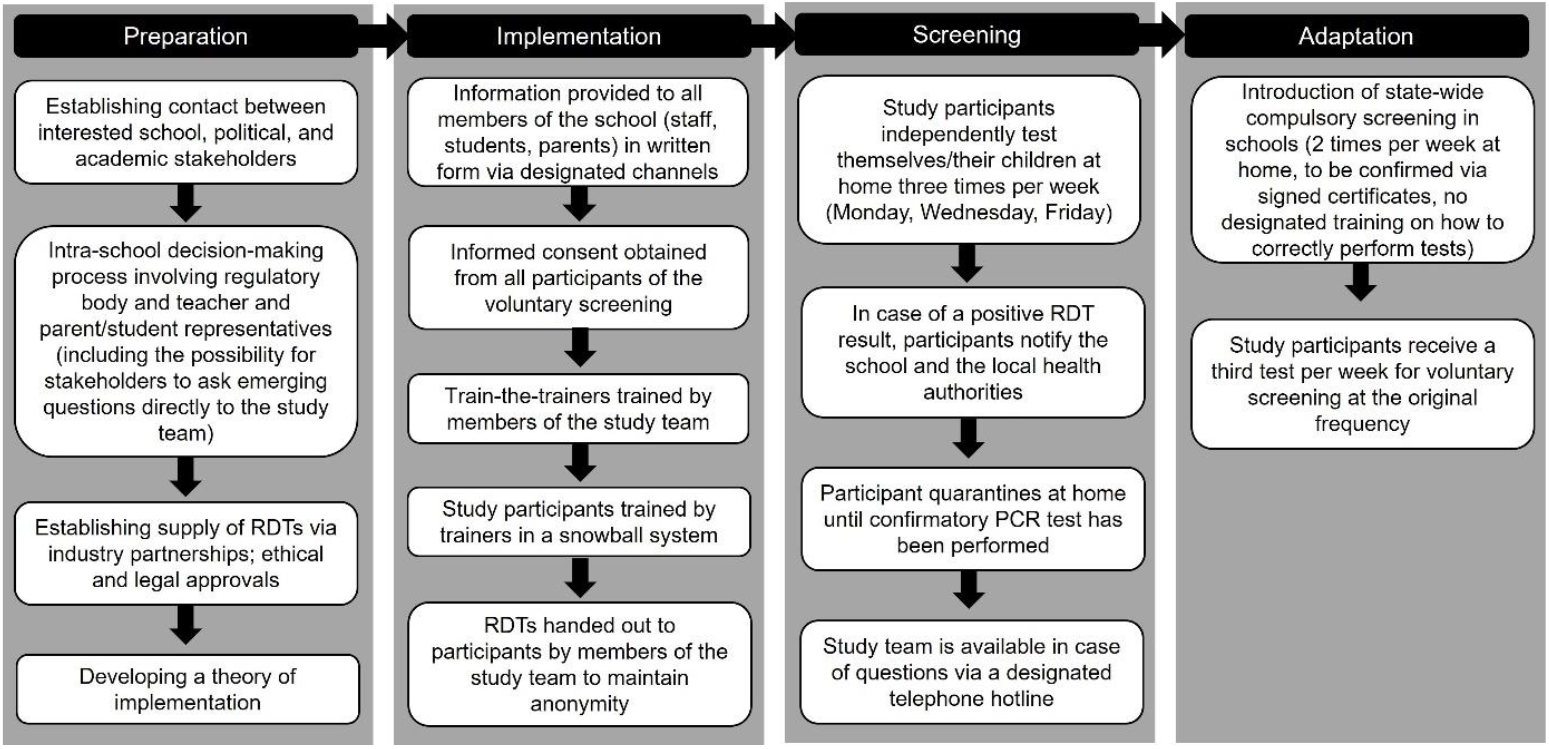
Implementation and theory of intervention of the RDT-based screening.

We trained members of the school staff and parents who volunteered on how to perform the test, and then trained others in a snowball system. We additionally set up telephone and email hotlines that could be contacted in case of screening-related questions. Additionally, the local health authority and local doctors were informed.

After four weeks of screening, compulsory testing was introduced for all schools in the German state of Baden-Württemberg.[12] The design of this state-wide screening was very similar to the study intervention, with the main difference being only two tests per week and parents having to confirm the test result to the school in writing. No training was offered in the context of the state-wide screening. Upon onset of the compulsory screening, all study participants were supplied with one test per week to supplement the two RDTs provided by the state to maintain the original screening frequency.

### Data collection and analysis

Quantitative (number of participants, number of tests handed out, and number of tests reported to be positive) and qualitative (in-depth interviews with children, parents, and school stakeholders) data were collected over the entire duration of the study; data collection and analysis procedures are outlined in Figure 3. All participants provided written informed consent separately for their participation in the screening and, if applicable, when they participated in the qualitative interview. The ethical review board at the Medical Faculty, Heidelberg University, Germany approved this study (S-141/2021). Further information on recruitment and data collection processes are reported in supplemental file 1. We followed COREQ-guidelines [13] to report our findings (see supplemental files 1 and 2).

**Figure 3.**
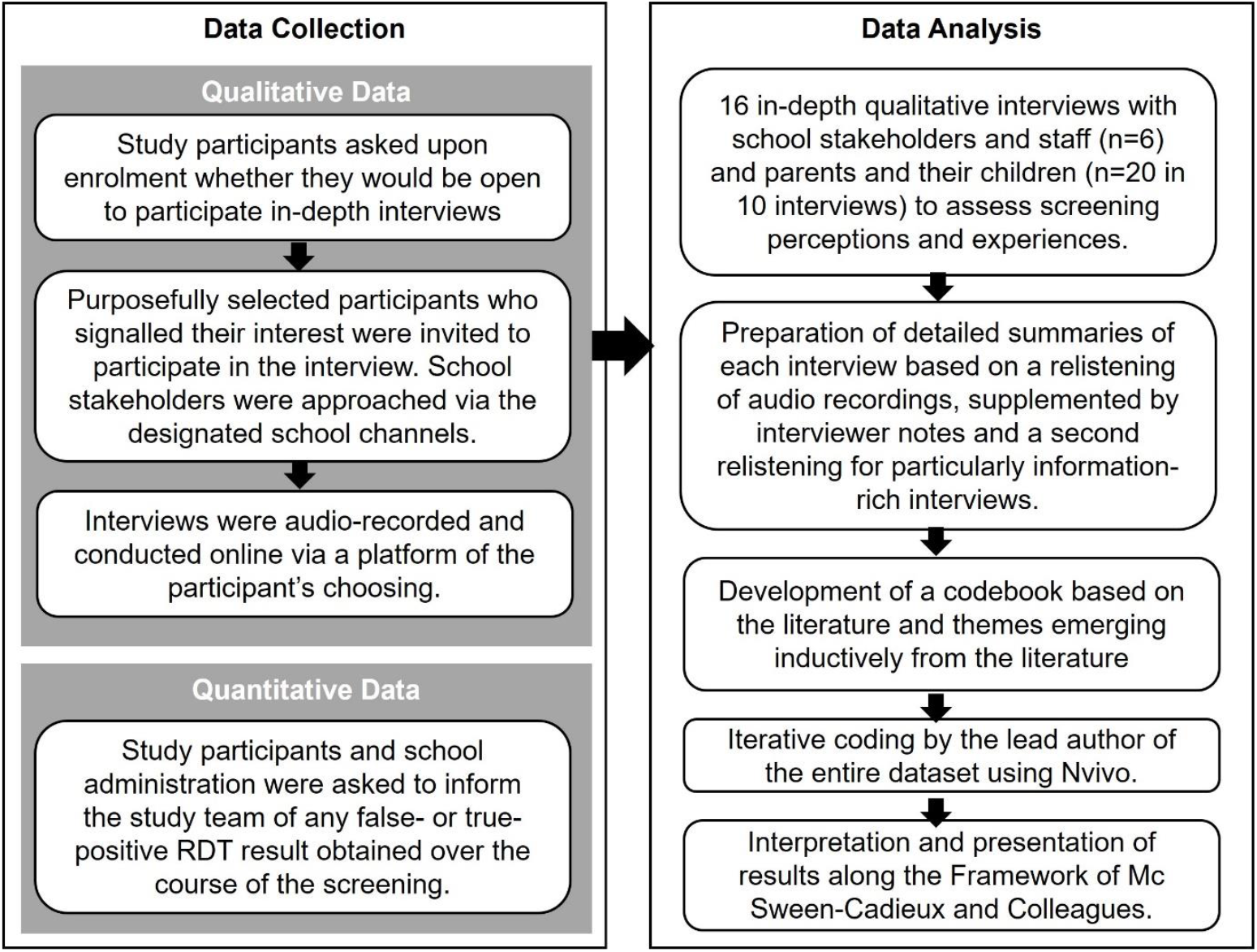
Data Collection and Analysis.

### Patient and public involvement

Members of the school administrative staff and parents of school children initiated contact with the study staff to express interest for developing a pilot project to assess the feasibility and acceptability of RDT-based screening at schools and were actively involved in the conceptualization and implementation of the study. School staff were not involved in participant recruitment and data collection to maintain anonymity. One co-author (NT) is parent to two school pupils and was responsible for initiating the study, and also participated in an interview as a key informant.

## RESULTS

### Study participants

A majority of school staff decided to participate in the voluntary screening (n=21 out of 34, 62%), as well as a majority of pupils and their parents (n=109 out of 186; 59%). The study lasted nine weeks. During this time, SARS-CoV-2 incidence in the region initially increased from 106.7 infections per 100,000 inhabitants per week (March 22) to 154.1 (April 27) before it fell to 54.3 by the end of the study-based screening (May 21).[14]

Over the course of the study, no study-related positive RDT-result (neither false-positive nor true-positive) was communicated to the school or the study team. After the onset of state-wide compulsory testing in schools, while the study-based screening was still in place, the school was notified of one case of SARS-CoV-2 in a student whose parents had self-reported a negative Ag-RDT one day prior. No further cases were reported.

### Implementation experiences, home-based testing

To highlight implementation processes and experiences, results are presented along the framework of McSween-Cadieux and colleagues (Table 1) [15] which combines the Consolidated Framework for Implementation Research [16] and the Ecological Framework.[17] The framework investigates factors influencing intervention implementation across six domains: intervention, individuals, support system, inner setting, outer setting, and the implementation process.

**Table 1.**
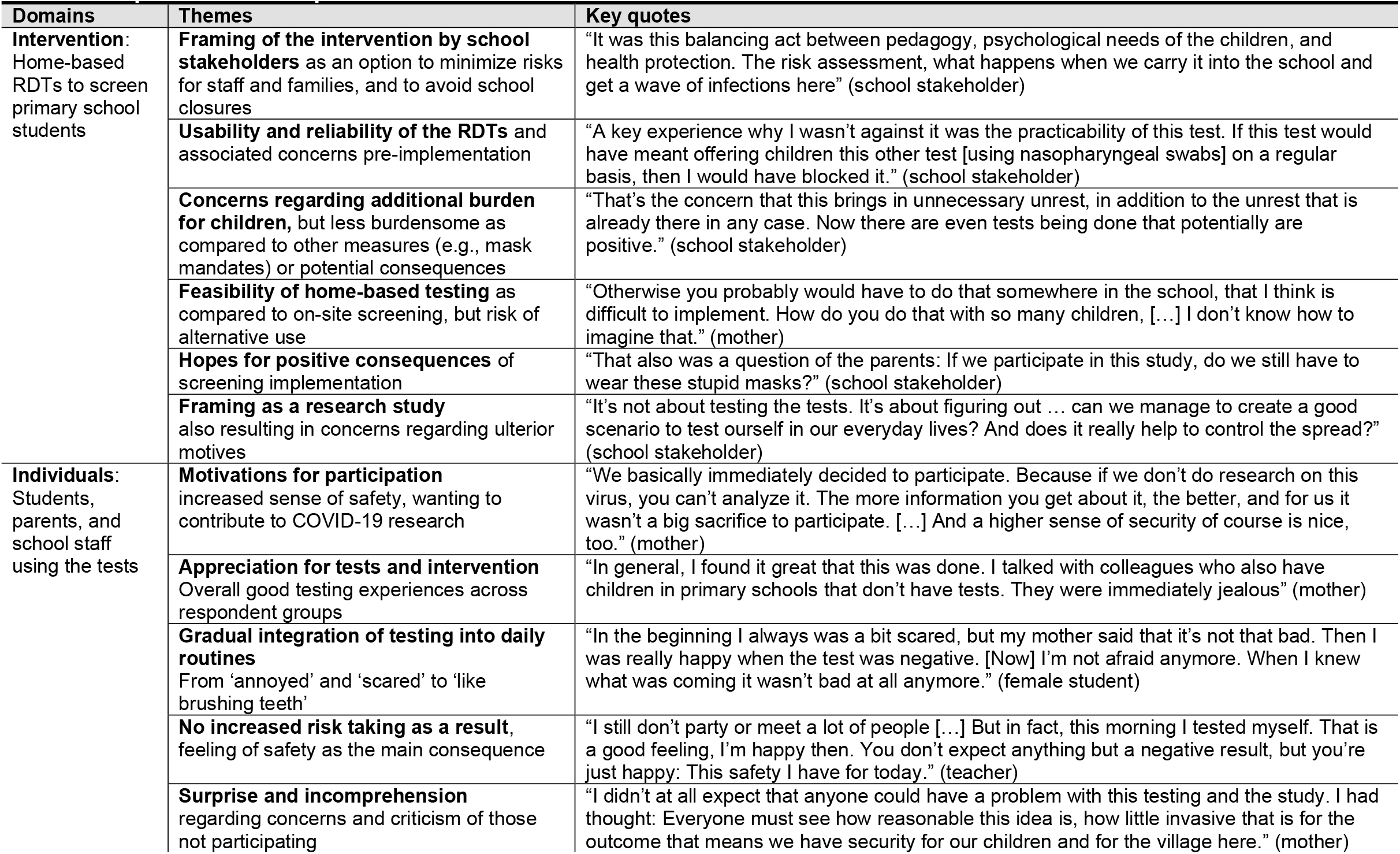
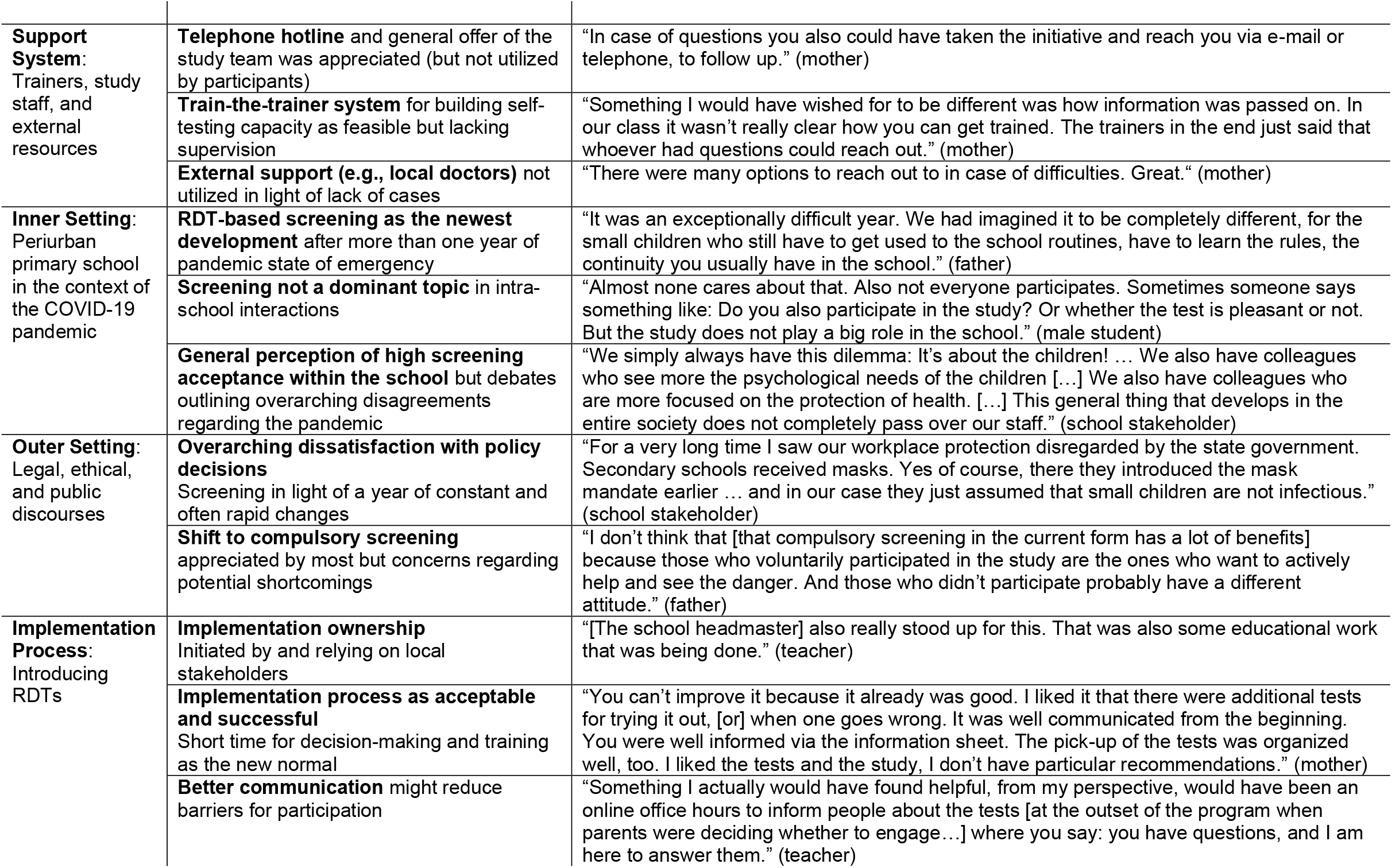
Implementation experiences across domains.

#### Intervention

School stakeholders highlighted their key motivations for exploring RDT-based screening as minimizing risks of secondary infections and school closures, as well as a hope that screening may lead to other positive consequences (e.g., repeal of mask mandates). Some participants voiced concerns regarding screening because it placed an unnecessary burden on children, especially in light of increasing communication at the time that children were not a driver of the pandemic and the perception that children’s physical and mental health was already strained enough by the pandemic. Consequently, a majority of stakeholders appreciated the newest generation of RDTs because they relied on anterio-nasal swabs, which were viewed as less burdensome for those performing the tests in general and children in particular.

A major point of debate entailed whether to conduct screening at home or on-site. School stakeholders and staff predominantly highlighted organizational and infrastructural barriers to school-based screening, including the strain on already limited teaching time, concerns regarding the psychological consequences of a student testing positive in school (including potential stigmatization by peers), and questions regarding teacher accountability. While participants generally acknowledged these concerns, several parents also discussed concerns that not everyone would conscientiously perform the tests at home. This was voiced when testing became compulsory, especially addressing families who initially had decided against study participation.

#### Individuals

An increased sense of safety was reported as the key motivation and consequence of testing across respondent groups for participation in the study and testing in general. Participants also reported a desire to contribute to COVID-19 research, thereby increasing the chance for a timely return to “a more normal school routine” (mother). The screening itself was generally appreciated, and a majority of participants described how initial reservations or “fear” (female student) regarding the tests were alleviated after the first few times, and the screening quickly was integrated into the morning routine “like brushing teeth” (mother). Children themselves described RDTs as being much less disrupting or burdensome compared to other measures encountered over the course of the pandemic, in particular compared to mask mandates in schools.

Several participants voiced incomprehension or “disappointment” (school stakeholder) regarding the number of families deciding against participation, or recounted frustration when interacting with screening hesitant parents or staff members. Several expressed disappointment regarding the limited consequences of participating in the screening and expected motivation and buy-in of others to increase once testing was seen as having consequences beyond a personal sense of security.

#### Support system

Participants appreciated offers made by the study team, including telephone and email hotlines, although neither was used during the course of the study. No participant reported interacting with complementary local resources (e.g., local health authority or local doctors). The experience of quick notification and confirmatory testing in light of a one positive results outside the study was seen as affirming that the support system in place would work.

Most participants appreciated the implemented train-the-trainer system and reported their interactions during the training as reassuring and empowering for when they performed the first RDT with their children, particularly when mistakes emerged during training. Participants saw themselves as being better prepared and able to assist others when statewide compulsory screening was implemented without prior training.

#### Inner setting

In light of prior experiences like school closures, challenges associated with remote learning, and quarantines, the intervention was perceived as less disruptive compared to other measures and associated with the hope for some continuity “at least until the summer break” (school stakeholder).

Both children and teachers reported the study-based screening to be only a side topic, if at all, in their interactions at school, although participating teachers recounted how sometimes children talked about their experiences in class or the reasons why their parents were against testing. In general, participants perceived screenings (both as part of the study and following the introduction of compulsory screening) as being highly accepted.

#### Outer setting

Participants stated that their support of the study-based screening represented an attempt to increase their own safety, which they felt had been neglected by elected authorities.

The subsequent introduction of compulsory screening therefore was appreciated by most participants, although concerns were voiced that a stricter control of testing fidelity than currently in place might be required, as not everyone was eager to comply.

#### Implementation process

The study was advocated by school stakeholders, and also relied on those stakeholders for successful implementation. This resulted in high level of stakeholder ownership, which was seen as particularly relevant for study buy-in across respondent groups. The broad buy-in was particularly important given a context marked by a highly emotionalized debate around COVID-19 control measures in schools.

Respondents generally appreciated the chosen implementation process. Although the information sheets, particularly the information sheet for children, and the communication by school stakeholders was appreciated, respondents expected study participation to further increase with additional events and opportunities for potential respondents to ask questions directly of the study team prior to making a decision about participation (which was only offered to the parents’ association and staff, though not all parents). Although the implemented training was highly appreciated, in a few instances, the snowball system did not work as envisioned, with information only being relayed verbally.

## DISCUSSION

This study outlined experiences implementing home-based RDTs for universal screening in a primary school setting. The screening was highly accepted among participants and viewed as feasible. Negative consequences were not observed (e.g., more risk-taking behavior). However, concerns surfaced regarding broad utility of screening when many individuals within a social setting may decline participation or not perform tests as advised. Participants expected screening acceptance and motivation to increase if the test was perceived to have consequences beyond a heightened sense of personal security. No case of SARS-CoV-2 was detected via the screening in the context of this study, and no clusters of infections indicated undetected cases.

To the best of our knowledge, this study is among the first to explore perceptions of test-based screening in a school setting. Our findings regarding the screening’s feasibility mirror outcomes of projects that implemented self-sampling for SARS-CoV-2 testing in school settings.[18, 19] However, this evidence stems from secondary schools [19] or from oral self-sampling.[18] The high acceptance of screening expressed by our participants mirrors qualitative evidence regarding the acceptance of broader COVID-19 prevention measures in schools in the UK.[7] We expand on this evidence by highlighting the acceptability and feasibility of home-based nasal sampling among primary school pupils.

The topic of large-scale RDT-based screening efforts in schools is emotionally highly charged in Germany, including lawsuits and homeschooling by parents who are fundamentally against SARS-CoV-2 testing for their children.[6] Our findings highlight that an emotionally charged intervention can be easily implemented if stakeholder buy-in and ownership is achieved through repeated explanations and demonstrations of the intervention. Also, our study demonstrated that testing was perceived as less burdensome to participants, including children, than more established measures, such as facial masks. Considering the exceptional burden faced by students, teachers, and parents in the pandemic,[20] and in light of increasing evidence regarding the impact of school closures on health and education [21, 22] and that children are unlikely to play a key role as drivers of the pandemic,[23] our results provide evidence that RDT-based screening is an acceptable and feasible way to facilitate in person teaching.

Beyond COVID-19, one other public health measure relying on self-testing in school settings in high-income countries entails screening for head lice. A study in primary schools in Australia aimed to assess the reliability of home-based screening for head lice, and only found a sensitivity of parental reports of 16%.[24] This suggests challenges of shifting testing from schools into the private realms, particularly in cases where a positive test result could be perceived as stigmatizing or as having consequences for short-term school access. While this concern was also voiced by participating parents and educators in our study, the participation of over 50% of staff and parents, probably reflects important distinctions between routine lice screening and self-testing amidst a viral pandemic.

This study provides timely and in-depth qualitative data, producing insights into the real-life discourse amidst rapidly changing regulations. The study site is representative for schools in peri-urban settings; research to date on health interventions at schools has predominantly focused on the urban context. However, our study also has limitations. First, only parents and students who had decided to voluntarily participate in the overarching screening program could be recruited for interviews; critical voices therefore might be underrepresented in the data on parents and their children. Additionally, as RDTs for SARS-CoV-2 have been introduced in Germany on a large scale in recent months, generalizability of our results to other countries where RDTs were less present in the public discourse might be limited.

RDT-based screening is an acceptable and easily scalable intervention to decrease risk of transmissions at schools and facilitate face-to-face teaching amidst a pandemic. Policymakers should ensure comprehensive capacity building for testing, fit-for-purpose training materials for all age levels and train-the-trainer programs to enable scale up of universal screening. Furthermore, consistent communication on regulations and readily available support networks (hotlines via phone or email) can reduce burden for school staff and families.

## Supporting information

Supplemental file 2 - COREQ checklist

## Data Availability

Considering the high public interest in research on COVID-19, qualitative data of participants who have indicated their agreement to this as part of the informed consent procedure can be shared with other researchers. However, to preserve the anonymity of respondents and considering the personal nature of qualitative data, requests will be considered on a case-by-case basis. Please contact the corresponding author.

## ACKNOWLEDGMENTS

We are grateful to all participants for their time and contribution.

## AUTHOR CONTRIBUTION

NT and CMD conceived of the study. JW, NT, SAM, and CMD conceptualized the study and data collection processes. JW, MS, and NT implemented the study with support from SAM and CMD. JW collected and analyzed the data, supported by all co-authors. JW drafted the manuscript. All co-authors contributed to data interpretation, edited the manuscript, and approved of the final manuscript.

