## Supplemental file 1 - methods for "“Like brushing teeth” – Implementation experiences with opt-in, at-home screening for SARS-CoV-2 among schoolchildren in Germany"

**PARTICIPANT RECRUITMENT AND DATA COLLECTION (Ctd.)**

The lead author (JW) who has graduate level education and several years of experience in conducting qualitative research conducted all interviews. He acknowledges that working in his country of origin and physically close to where he lives and works could result in biases, especially in light of being affected by the same COVID-19 associated restrictions and policies as respondents.

Interviews were scheduled via email and conducted on a videocall platform of the participant’s choosing. We contacted a total of 31 parents who had signaled potential interest in participating in an interview and ultimately enrolled 10. Reasons for not participating in the interviews in all cases were associated with scheduling difficulties and the high workload of managing homeschooling for primary school children while working from home oneself. Prior to each interview, the interviewer described content of the interview and the own interest in the topic at hand and provided participants the opportunity to ask any open questions. To the best of the interviewer’s knowledge, no third party was present in the room with respondents during the online interviews.

The semi-structured interview guide, including questions and further probes, was developed based on the literature and the study team’s previous experience on conducting qualitative interviews on SARS-CoV-2 RDTs in Germany. The interviews on average lasted 45 minutes (range 24 to 74 minutes) and concluded once saturation was reached (for interviews with parents and their children) or all respondents expressing interest to participate were interviewed (for school stakeholders and staff).
